# DBS-based Biomarkers of Neurodegeneration predict Cognitive Decline and Dementia Seven Years Ahead in a Large Population-Based Sample

**DOI:** 10.64898/2026.01.07.26343614

**Authors:** Axel Börsch-Supan, Nis Borbye-Lorenzen, Yacila Deza-Lougovski, Salima Douhou, Thorsten Kneip, Marcela C. Otero, Yuri Pettinicchi, Martina Börsch-Supan

## Abstract

**INTRODUCTION:** It is unknown whether biomarkers of neurodegeneration collected from dried blood spots (DBS) in large-scale population settings are useful in predicting cognitive decline many years later.

**METHODS:** In 2015, we collected DBS of 13,679 individuals aged 58 and older. All DBS were assayed for ApoE4 protein, and a smaller subsample for pTau217, GFAP, and NfL. In 2022, we obtained detailed cognition measures for 6,523 respondents. Regression analyses tested the likelihood of cognitive impairment as a function of biomarker levels.

**RESULTS:** Respondents with ApoE4 detection have worse cognitive performance seven years later as measured by six different cognitive performance measures (p<0.001). The combination of all four biomarkers is a statistically significant predictor for five cognitive performance measures, with pTau217 having the most systematic association.

**DISCUSSION:** DBS-based biomarkers of neurodegeneration provide a cost-efficient and scalable early warning signal enabling preventive measures against AD/ADRD before the onset of serious symptoms.

## 1. Background

This study provides evidence that biomarkers of neurodegeneration, such as the ApoE4 protein, pTau217, GFAP, and NfL, which can be measured in dried blood spots (DBS), are useful in predicting cognitive decline and dementia many years later in large-scale population settings. The significance of this study is threefold. First, we collected DBS from a representative population sample under normal survey conditions in the community, outside of a clinical setting and with minimal invasion. This is different from studies such as Wu et al.^1^, who employ plasma samples drawn from hospital patients. Second, we use easily collected DBS rather than venous blood, such as Faul et al.^2^ and Kim et al.^3^. Third, we study predictions seven years ahead. This is different from studies such as Deza-Lougovski et al.^4^, Faul et al.^2^ and Wu et al.^1^ who show contemporaneous associations. Our study provides therefore a cost-efficient and scalable early warning signal enabling preventive measures against AD/ADRD in large populations long before the onset of serious symptoms.

Early detection is important since dementia is currently without cure. Early prevention measures, such as life-style changes recommended by the Lancet Commission^5^, are therefore the key to reducing the dementia burden. This burden is large. According to the Global Burden of Disease (GBD) 2019 report, AD/ADRD is the 4th leading cause of disease burden among people aged 75 years and over, and accounts for 5.6% of years lived with disability. AD is the 6th leading cause of death in the US^6^. GBD estimates 57.4 million people with dementia in 2019, the numbers nearly doubling every 20 years, to 83.2 million in 2030 and 152.8 million in 2050^7^.

The sample of this study is unique because it is based on a large representative population study of individuals aged 50 years and older in Europe and Israel (SHARE, the Survey of Health, Ageing and Retirement in Europe), which collected blood in 2015 as part of its Wave 6, and measured cognitive performance seven years later as part of its Wave 9. Specifically, SHARE collected DBS in 2015 in twelve countries (Belgium, Denmark, Estonia, France, Germany, Greece, Israel, Italy, Spain, Slovenia, Sweden, and Switzerland). In 2018, 13,679 DBS samples from all twelve countries were analyzed for the ApoE4 protein. In 2025, 3103 DBS samples with ApoE4 information from three countries (Denmark, Germany and Sweden) were also analyzed for pTau217, GFAP and NfL. We obtained multiple indicators of cognitive performance in 2022, including three basic cognitive performance measures (immediate and delayed word recall, orientation in time) and three cognition summary scores used in the literature (HCAP-validated score^8^; Global cognition score^4^; Langa-Weir score^9^).

We find that a level of the ApoE4 protein that indicates the presence of at least one *APOE ε4* allele is statistically significantly associated with worse cognitive performance seven years later as measured by all three basic cognitive performance measures and all three cognition summary scores. We also find that pTau217 is significantly associated with worse cognitive performance as measured by the immediate word recall test and the HCAP-validated and Langa-Weir cognition summary scores. GFAP is significantly associated with a lower ability to orient oneself in time, while we find no convincingly significant associations for NfL. Notably, the associations with cognitive performance are statistically significant although we systematically account for potential confounding variables. Actually, strength and significance of the associations increase as we sequentially add more confounders such as demographics, education and comorbidities. This suggests that the relationship between the biomarker levels and cognitive decline is direct and not merely a spurious correlation driven by other factors. We also find that the associations are robust against sample attrition between Waves 6 and 9 of the survey. We conclude that the four biomarkers of neurodegeneration collected from DBS in a representative population sample and under normal survey conditions are useful and efficient predictors of cognitive impairment many years later, providing a foundation for early prevention measures to reduce the dementia burden.

## 2. Methods

We describe the study population, define our measures of cognitive performance and neurodegenerative biomarkers, the set of other modifiable and non-modifiable risk and protective factors for cognitive impairment to be controlled for, and present our statistical methodology.

### 2.1 Study Population

This study utilizes data from the Survey of Health, Ageing and Retirement in Europe (SHARE), a large multidisciplinary, longitudinal survey representative of individuals aged 50 and older across 28 European countries, based on multi-stage probability sampling from administrative registers and similar sources^10^. To complement self-reported health data, dried blood spot (DBS) samples were collected during SHARE Wave 6 (2015) from 13,679 individuals aged 58 and older across 12 participating countries (Israel, Spain, Greece, Italy, Switzerland, France, Slovenia, Belgium, Germany, Denmark, Estonia, and Sweden). These DBS samples were analyzed for various biomarkers, including ApoE4 protein levels, with valid measurements obtained for 13,102 respondents^11^.

To examine prospective associations, we selected individuals who participated in both Wave 6 (2015) and Wave 9 (2022), were aged 65 or older in 2022, had valid ApoE4 biomarker data in Wave 6 and valid cognition data in Wave 9. The final analytic sample for this prospective analysis comprised 6,523 respondents (58.6% female). To adjust for potential bias related to non-response and selective participation, potentially related to cognitive impairment, all population statistics were computed using weights that accounted for the probability of participating in Wave 9 conditional on participation in the biomarker collection of Wave 6. These weights were derived using inverse probability weighting based on a logistic regression model. The model predicted Wave 9 participation using age (in 5-year bands), sex, educational attainment, and country of residence, all measured in Wave 6. This specification is also used to test for robustness of the associations with respect to sample attrition.

### 2.2 Biomarkers of neurodegeneration

The DBS samples were analyzed for seven conventional biomarkers^11^, a set of cytokines, the ApoE4 protein, pTau217, GFAP, and NfL.

#### ApoE4 Protein

Apolipoprotein E4 (ApoE4) levels were quantified in 2018 alongside a set of cytokines using a 10-plex immunoassay on Meso Scale Discovery (MSD) plates. The assay procedure involved extracting the protein from the DBS cards, calibrating the measurements against recombinant human Apoe4 protein, and detecting the specific isoform using tailored antibodies. Methodological details, including assay validation and quality control protocols, are documented^12^. The authors then validated the results of the immunoassay with the corresponding genetic information, namely the presence of at least one *APOE ε4* allele, demonstrating 97% sensitivity and 98% specificity. This study established a validated threshold of 30,000 pg/mL for specific binding, which reliably indicates the presence of at least one *APOE* ε4 allele in genotyped individuals. The study also showed that the ApoE4 protein measure is robust to survey conditions, such as ambient temperature, spot size, drying time, shipping time to the lab, and packaging mode. Deza-Lougovski et al.^4^ showed that the presence of the ApoE4 protein was associated with worse cognitive performance in the SHARE sample of respondents aged 50 and older. For the present analysis, ApoE4 protein levels were dichotomized based on this threshold, distinguishing between individuals with and without detected binding. As a shorthand, we will denote this as “ApoE4 detection”.

#### pTau217, GFAP and NfL

Phosphorylated Tau at threonine 217 (pTau217), glial fibrillary acidic protein (GFAP) and neurofilament light chain (NfL) were analyzed much later in 2025 from remaining DBS from the countries Denmark, Germany and Sweden, where we had permission to store them longer and had more material left relative to other countries. Criteria to be analyzed included sufficient blood material for the assay on the remaining DBS sample (an area allowing to collect a subsample of two punches of 3.2 mm in diameter) and the previous determination of ApoE4 status for this respondent. GFAP and NfL were analyzed jointly with Tau (total) using the S-PLEX Neurology Panel 1, MSD’s ultrasensitive assay platform that reduces the lower limit of detection for immunoassays, whereas pTau217 was analyzed separately in a single-analyte format (MSD’s S-PLEX Human Tau (pTau217) Kit). Only 0.2% of the analytical sample fell below the detection limit for pTau217, 0.7% for GFAP and 8.1% for NfL. Intra(inter)assay CV was 22.8% (19.3%) for pTau217, 15.0% (13.4%) for GFAP and 18.7% (14.3%) for NfL.

pTau217 and NfL are robust to survey conditions, while GFAP was sensitive to ambient temperature, drying time and whether the shipping bag was open or closed (Supplementary Section S1). Huber et al.^13^ report significant changes of GFAP levels in dried plasma spots over time and at elevated temperature and find NfL and pTau181 stable at longer storage and higher temperatures. In our statistical analyses, we therefore correct for these survey conditions.

### 2.3 Measures of Cognitive Impairment

The SHARE data include a large set of cognitive impairment indicators derived from performance tests administered as part of the SHARE survey, including among others verbal episodic memory (immediate and delayed recall), temporal orientation, verbal fluency (animal naming), and numeracy (serial subtraction). A comprehensive description of the individual cognitive tests and the procedures used to compute the cognitive impairment indicators is provided in Supplementary Section S2.6. We also condensed these tests to three recently developed summary measures of cognitive impairment: The Global Cognitive Impairment score (Gscore), the Langa-Weir scale (Lscore), and a probability-based measure (Hscore) validated by an in-depth cognitive assessment using the Harmonized Cognitive Assessment Protocol^14^. To be comparable, the three summary scores are standardized to mean zero and standard deviation one.

#### Global Cognitive Impairment score (Gscore)

Gscore is constructed by averaging standardized (Z-scored) cognitive test scores of five indicators (verbal episodic memory (immediate and delayed recall), temporal orientation, verbal fluency (animals), and numeracy (serial subtractions). For detailed methodological information, refer to ^4^.

#### Langa-Weir (Lscore)

The Langa-Weir classification provides both a continuous summary score (range between 0 and 27) that adds the outcome of four cognition tests (immediate and delayed recall, backwards counting and serial subtractions) and a categorical classification of cognitive status that assigns respondents to one of three categories: Normal, Cognitively Impaired but Not Demented (CIND), or Demented. The thresholds for this classification have been validated in large-scale population aging studies^9,15,16,17,18^. In this study, we use the continuous summary score and standardize it to mean zero and standard deviation one.

#### HCAP-refined Cognitive Impairment score (Hscore)

The Harmonized Cognitive Assessment Protocol (HCAP) was developed by Langa et al.^14^ to enable international comparisons of cognitive performance. It includes some 27 cognitive performance tests that can be condensed to various summary scores. SHARE administered HCAP in Wave 9 (2022) to a subsample of 2,685 individuals in Germany, Italy, France, Denmark and the Czech Republic. Otero et al.^19^ describe how these in-depth cognitive tests were condensed to five dimensions using confirmatory factor analysis. With the help of classification using standard diagnostic criteria^20^, these five dimensions were used to classify the respondents into three cognitive performance states: Normal, Mild Cognitive Impairment (MCI) and Dementia. Employing a regression-based approach, Börsch-Supan et al.^8^ use the results from the HCAP subsample to validate the cognition measurement in the entire SHARE Wave 9 sample. The regression outcome is the probability of suffering from cognitive impairment (MCI or dementia), which is standardized to mean zero and standard deviation one.

### 2.4 Other risk and protective factors

Key aim of this study is to relate the biomarkers of neurodegeneration based on DBS collected in 2015 to measures of cognitive performance obtained seven years later, correcting for other modifiable and non-modifiable risk factors sourced from SHARE. They include demographics, education and comorbidities. The biomarkers are considered proxies for inherent biological susceptibility, while education and comorbidities are viewed as modifiable risk factors potentially offering cognitive protection. This dual perspective underscores the interplay between intrinsic (genetic) and extrinsic (environmental) influences on the trajectory of cognitive aging.

#### Education

Educational attainment was assessed using the International Standard Classification of Education (ISCED-97), a harmonized framework developed by UNESCO to facilitate meaningful cross-national comparisons of educational systems. ISCED-97 classifies educational levels into seven categories, ranging from pre-primary education (level 0) to advanced university degrees (level 6). For the purpose of this analysis, these education levels were coded according to the ISCED-97 scale and treated as dummy variables. This approach allowed for the examination of potential non-linear relationships between different levels of educational attainment and cognitive outcomes.

#### Comorbidities

Other potentially influential and modifiable risk factors included comorbidities. Functional status was assessed using the Activity of Daily Living (ADL) score (ranging from 0 to 6) and the Instrumental Activities of Daily Living (IADL) score (ranging from 0 to 9). Additionally, a cardiovascular multimorbidity index (CVR), as defined in ^21^, body mass index (BMI), and the presence of depressive symptoms (EURO-D^22^) were included as well as indicators for the prevalence of high cholesterol, hypertension and affective disorders.

#### Non-modifiable risk factors

We also control for demographic characteristics (age and sex) and survey-technical variables. To mitigate the potential influence of repeated cognitive assessments, the number of prior SHARE wave participations was included as a covariate, acting as a proxy for cognitive practice effects. Finally, a series of country indicator variables were incorporated to control for national-level differences not captured by the other explanatory variables, such as variations in access to healthcare systems.

Detailed information regarding the specific SHARE interview items, coding procedures, and scoring algorithms for these covariates can be found in Supplementary Section S2.

### 2.5 Statistical Methods

The main analyses use linear regression methods with measures of cognitive performance as dependent variables and biomarkers of neurodegeneration and other risk factor as explanatory variables. Statistical significance was defined as a p-value less than 0.05.

Missing data for the regressors were relatively limited. Specifically, 103 observations (1.6% of the analytical sample) had missing values for the EURO-D depression scale, and 144 observations (2.2% of the analytical sample) had missing BMI values. To mitigate the impact of missingness in the EURO-D variable, a missingness indicator (dummy variable) was included in the models to retain these observations. For BMI, we utilized imputed values as released by the SHARE project. The number of observations with missing values for other covariates and the corresponding handling procedures are detailed in Supplementary Section S2.8. Notably, we observed a slightly higher rate of missingness among cognitively impaired respondents. Descriptive statistics comparing the dropped observations to the analytical sample with respect to cognitive status are also provided in that supplement.

## 3. Results

The results section is organized into five parts. First, we show descriptive statistics of cognitive performance as dependent variable and biomarkers of neurodegeneration and other risk factors as independent variables. Second, we examine the seven-year predictive power of ApoE4 detection in the sample of 12 countries. Third, we do the same for pTau217, GFAP and NfL each separately in the much smaller sample of three countries. Fourth, we combine all biomarkers in a joint analysis. Finally, we perform robustness analyses, especially with respect to sample attrition between Waves 6 and 9.

### 3.1 Descriptive analysis

The analytic sample of the 12 countries with ApoE4 in Wave 6 and detailed cognition data in Wave 9 comprises 6,523 observations, representing a diverse group of individuals in older adulthood across cognitive, health, demographic, and lifestyle domains. The analytic sample of the three countries with data on pTau217, GFAP and NfL is substantially smaller with 1,282 observations. Supplementary Table S3 provides means and standard deviations of all variables used in this study, based on the sample of 6,523 observations. The majority of participants were female (58.6%). The age distribution was: 58–64 (31.3%), 65–69 (19.5%), 70–74 (15.0%), 75–79 (13.7%), 80-84 (11.7%), 85–89 (6.2%), and 90+ (2.8%). In terms of educational attainment, the largest groups held either a higher secondary school degree (33.3%) or a college degree (28.9%).

The dependent variables in our study are three often used measures of cognitive performance (immediate and delayed word recall, temporal orientation) and three summary indices used to classify cognitive impairment. Immediate word recall yields a mean of 5.1 words (SD=1.8); delayed word recall 3.7 words (SD=2.1). Temporal orientation fails for 16% of the sample. Using their respective thresholds, the summary indices show a prevalence of mild or severe cognitive impairment of 5.4% using the general cognition Gscore, 13.2% with the Langa-Weir Lscore, and 9.0% using the HCAP-validated Hscore. Börsch-Supan et al.^8^ present the prevalence of severe cognitive impairment across countries based on Hscore. It ranges from 5% in Sweden to 22% in Spain. The Lscore exhibits an even larger international variation from 2% severely impaired to 29%, with a very similar ranking by countries. The three cognition summary measures are highly correlated since they have common underlying cognition indicators. Hscore and Gscore exhibit a correlation of r=0.77, Hscore and Lscore of r=0.67, and Gscore and Lscore of r=0.93. These correlations imply that the three summary measures capture similar aspects of overall cognitive ability.

Explanatory variables are the four biomarkers of neurodegeneration. Genetic risk, as indicated by ApoE4 detection, was observed in 26.2% of the sample. Among the respondents with ApoE4 detected, the share of cognitively impaired individuals is higher (13.1%) than among those without ApoE4 (11.7%) as measured by Hscore. This is an increase by 11.6%. This is the raw effect to which the regression estimates that control for other potentially correlated risk factors have to be compared. pTau217 has a mean value of 2.08 pg/mL (SD=1.8), GFAP 1.35 pg/mL (SD=1.0) and NfL 3.33 pg/mL (SD=4.3).

Figure 1 divides the study sample into four groups defined by the combination of ApoE4 protein concentration (x-axis, low vs. high) and the estimated probability of being cognitively impaired that defines the Hscore (y-axis, normal vs. impaired) and its dichotomized variant, denoted by HCI. The vertical dashed line at 30,000 pg/mL indicates the threshold above which specific ApoE4 binding is detected. Values to the left of this cutoff reflect undetectable or non-specific binding. Observations classified as cognitively normal are marked by green diamonds, while those identified as cognitively impaired are shown as red circles.

**Figure 1:**
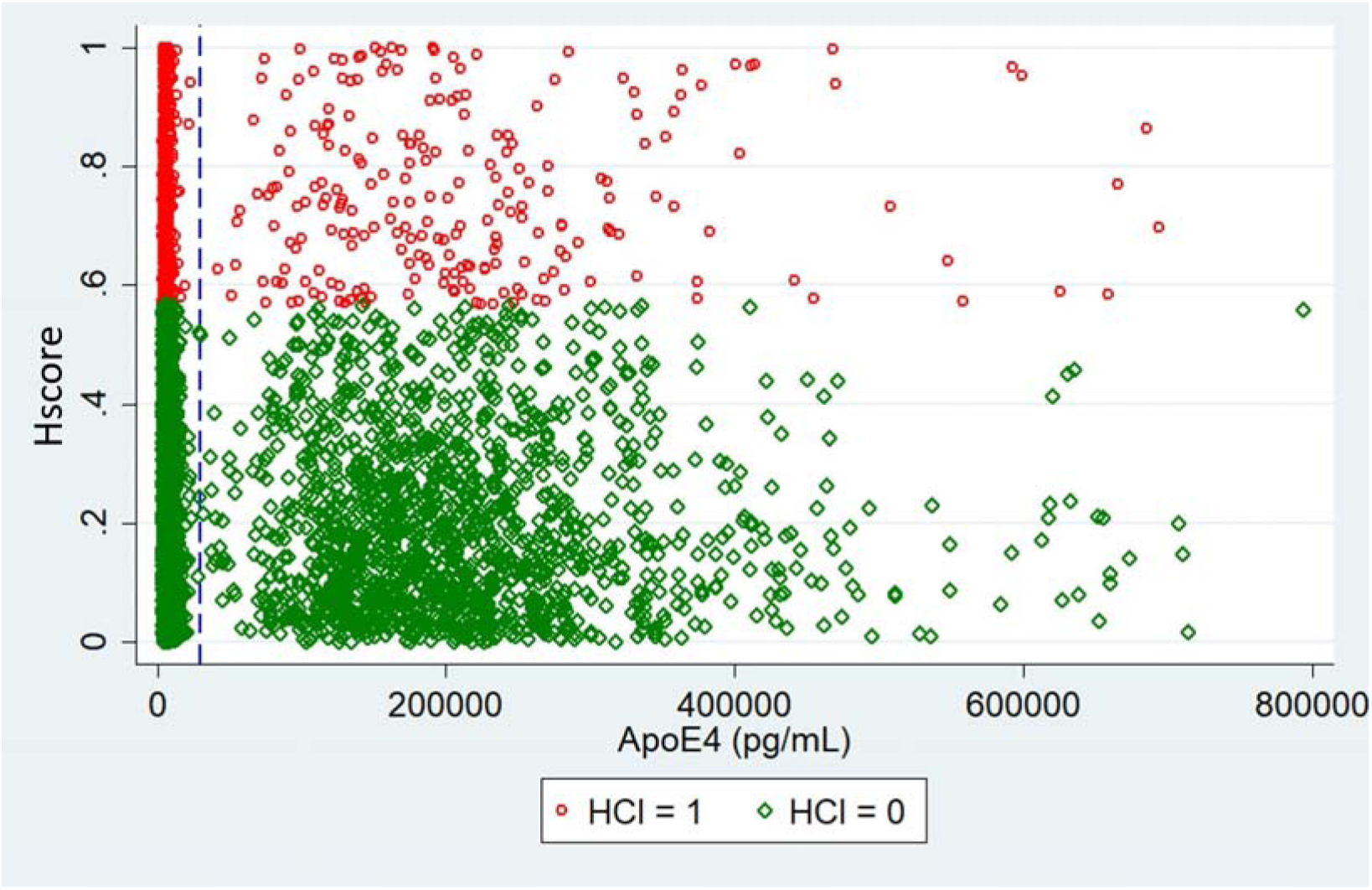
**ApoE4 protein level and HCAP-validated cognitive impairment** The horizontal axis displays the ApoE4 protein concentration. 30,000 pg/mL indicates the threshold above which specific ApoE4 binding is detected. The vertical axis shows the estimated probability of being cognitively impaired based on Hscore and its dichotomized variant HCI. Observations classified as cognitively normal (cognitively impaired) are marked by green diamonds (red circles).

Comorbidities were measured at baseline. The average body-mass index (BMI) was 26.8 (SD= 4.8), just above the threshold of overweight status. On average, participants contributed data across 4.9 waves (SD = 1.2). Functional limitations showed large variability, with an average of 0.23 (SD = 0.78) for activities of daily living (ADL) and 0.55 (SD = 1.43) for instrumental activities of daily living (IADL). Affective symptoms were rare (mean = 0.06), and the cardiovascular risk (CVR) score averaged 0.33 (SD = 0.57). The prevalence of high cholesterol and hypertension was 31% and 56%, respectively. Most participants reported to be free of depressive symptoms (70%).

### 3.2 Seven-year predictive power of ApoE4 detection

We first investigate how well ApoE4 predicts cognitive performance seven years later. Table 1 shows the results.

**Table 1:** Predictive power of ApoE4 measured seven years earlier.

| Dependent variable | Explanatory variable is ApoE4_detected |  |  |  |  |
| --- | --- | --- | --- | --- | --- |
|  | Coeff | StdErr | p-value | R-squared | Nobs |
| **immword** | -0.143 | 0.042 | 0.001 | 0.303 | 6495 |
| **delword** | -0.235 | 0.052 | 0.000 | 0.279 | 6505 |
| **orientt** | -0.101 | 0.021 | 0.000 | 0.320 | 6660 |
| **Hscore** | -0.069 | 0.018 | 0.000 | 0.579 | 6470 |
| **Gscore** | -0.089 | 0.021 | 0.000 | 0.419 | 5942 |
| **Lscore** | -0.094 | 0.022 | 0.000 | 0.391 | 6521 |

**Notes:** The dependent variables are the six cognition measures in the first column. Besides the detection of ApoE4, the regressions include the following covariates: **Demographic** variables include age categories and sex. **Education** variables include ISCED-1997 education categories. **Comorbidity** variables include body mass index, EURO-D depression scale, ADL and IADL scores, CVR score, and indicators for affective symptoms, cholesterol and hypertension. Also included are country indicators and the number of waves participated.

Respondents with ApoE4 detected show statistically significant negative associations with all three basic measures of cognitive performance (immediate word recall, delayed word recall, temporal orientation). The same holds for all three summary indicators of cognitive performance (Hscore, Gscore, Lscore). In addition, these regressions deliver a good prediction accuracy, especially Hscore with an R-square of 0.58.

However, the effect sizes are small relative to the standard deviations of the cognition measures. Having ApoE4 detected reduces the performance in the immediate word recall test by 0.14 words and in the delayed word recall test by 0.24 words. Respondents with ApoE4 detected fail the temporal orientation test 10% more frequently. Since the summary scores are standardized, respondents with ApoE4 detected obtain cognition scores that are 0.07, 0.09 and 0.09 standard deviations lower for Hscore, Gscore and Lscore, respectively.

The associations with cognitive performance are statistically significant although we account for a large number of potentially confounding variables. Table 2 shows that strength and significance of the associations increase as we sequentially add more confounders such as demographics, education, comorbidities, and country effects. This suggests that the relationship between the biomarker levels and cognitive impairment is direct and not merely a spurious correlation driven by other factors.

**Table 2:** Robustness with respect to included covariates.

|  | **Coeff** | **StdErr** | **p-value** | **Coeff** | **StdErr** | **p-value** | **Coeff** | **StdErr** | **p-value** | **Coeff** | **StdErr** | **p-value** |
| --- | --- | --- | --- | --- | --- | --- | --- | --- | --- | --- | --- | --- |
| **Hscore** | -0.068 | 0.026 | 0.009 | -0.076 | 0.026 | 0.003 | -0.061 | 0.019 | 0.001 | -0.069 | 0.018 | 0.000 |
| **Gscore** | -0.062 | 0.025 | 0.015 | -0.081 | 0.023 | 0.001 | -0.073 | 0.022 | 0.001 | -0.089 | 0.021 | 0.000 |
| **Lscore** | -0.074 | 0.025 | 0.003 | -0.091 | 0.024 | 0.000 | -0.081 | 0.022 | 0.000 | -0.094 | 0.022 | 0.000 |
| Demographics | included |  |  | included |  |  | included |  |  | included |  |  |
| Education |  |  |  | included |  |  | included |  |  | included |  |  |
| Comorbidities |  |  |  |  |  |  | included |  |  | included |  |  |
| Countries |  |  |  |  |  |  |  |  |  | included |  |  |

**Notes:** The dependent variables are the three summary cognition measures in the first column. Besides the detection of ApoE4, the regressions include the following covariates: **Demographics** includes age categories and sex. **Education** includes ISCED-1997 education categories. **Comorbidities** includes body mass index, EURO-D depression scale, ADL and IADL scores, CVR score, and indicators for affective symptoms, cholesterol and hypertension. **Countries** includes indicators for each country.

These other factors are shown in Supplemental Section S4, taking the association between ApoE4 and Hscore as an example. Age and education are highly significant and have the expected direction: older respondents have a worse cognitive performance, and respondents with higher secondary education or some college, a better one. Comorbidities do not play a large role except for BMI and especially IADLs.

### 3.3 Seven-year predictive power of pTau217, GFAP and NfL

We now turn to pTau217, GFAP and NfL that were later analyzed for the much smaller sample of Denmark, Germany and Sweden. Table 3 shows the predictive power of pTau217 for the six measures of cognitive performance seven years later.

**Table 3:** Predictive power of pTau217 measured seven years earlier.

| Dependent variable | Explanatory variable is pTau217 |  |  |  |  |
| --- | --- | --- | --- | --- | --- |
|  | Coeff | StdErr | p-value | R-squared | Nobs |
| **immword** | -0.057 | 0.027 | 0.037 | 0.242 | 1205 |
| **delword** | 0.000 | 0.000 | 0.011 | 0.239 | 1204 |
| **orientt** | 0.001 | 0.022 | 0.959 | 0.272 | 1225 |
| **Hscore** | -0.032 | 0.016 | 0.040 | 0.402 | 1203 |
| **Gscore** | -0.039 | 0.025 | 0.116 | 0.321 | 1214 |
| **Lscore** | -0.060 | 0.025 | 0.014 | 0.292 | 1204 |

**Notes:** The dependent variables are the six cognition measures in the first column. Besides pTau217, the regressions include the following covariates: **Demographic** variables include age categories and sex. **Education** variables include ISCED-1997 education categories. **Comorbidity** variables include body mass index, EURO-D depression scale, ADL and IADL scores, CVR score, and indicators for affective symptoms, cholesterol and hypertension. Also included are country indicators and the number of waves participated.

Respondents with elevated pTau217 levels do statistically significantly worse in the immediate word recall test seven years later. There is less of a systematic association between pTau217 and the two other basic measures of cognitive performance (delayed word recall, temporal orientation). However, the summary indicators show a clear pattern: respondents with elevated pTau217 levels have a worse cognitive performance seven years later. The effect sizes are of a similar order of magnitude as for ApoE4.

Table 4 shows very systematic associations between GFAP and the six cognitive performance measures although the statistical significance is weak due to the small sample size. Elevated GFAP level reduce cognitive performance seven years later.

**Table 4:** Predictive power of GFAP measured seven years earlier.

| Dependent | Explanatory variable is GFAP |  |  |  |  |
| --- | --- | --- | --- | --- | --- |
| Variable | Coeff | StdErr | p-value | R-squared | Nobs |
| **Immword** | -0.028 | 0.030 | 0.340 | 0.238 | 1262 |
| **Delword** | -0.060 | 0.032 | 0.062 | 0.229 | 1261 |
| **Orientt** | -0.066 | 0.023 | 0.004 | 0.267 | 1282 |
| **Hscore** | -0.030 | 0.018 | 0.099 | 0.390 | 1259 |
| **Gscore** | -0.059 | 0.027 | 0.028 | 0.313 | 1271 |
| **Lscore** | -0.046 | 0.027 | 0.086 | 0.282 | 1261 |

**Notes:** The dependent variables are the six cognition measures in the first column. Besides GFAP, the regressions include the following covariates: **Demographic** variables include age categories and sex. **Education** variables include ISCED-1997 education categories. **Comorbidity** variables include body mass index, EURO-D depression scale, ADL and IADL scores, CVR score, and indicators for affective symptoms, cholesterol and hypertension. Also included are country indicators and the number of waves participated.

For the NfL biomarker, we find the same pattern in Table 5, but no association is statistically significant except – although very weakly – between the Langa-Weir score and NfL.

**Table 5.**
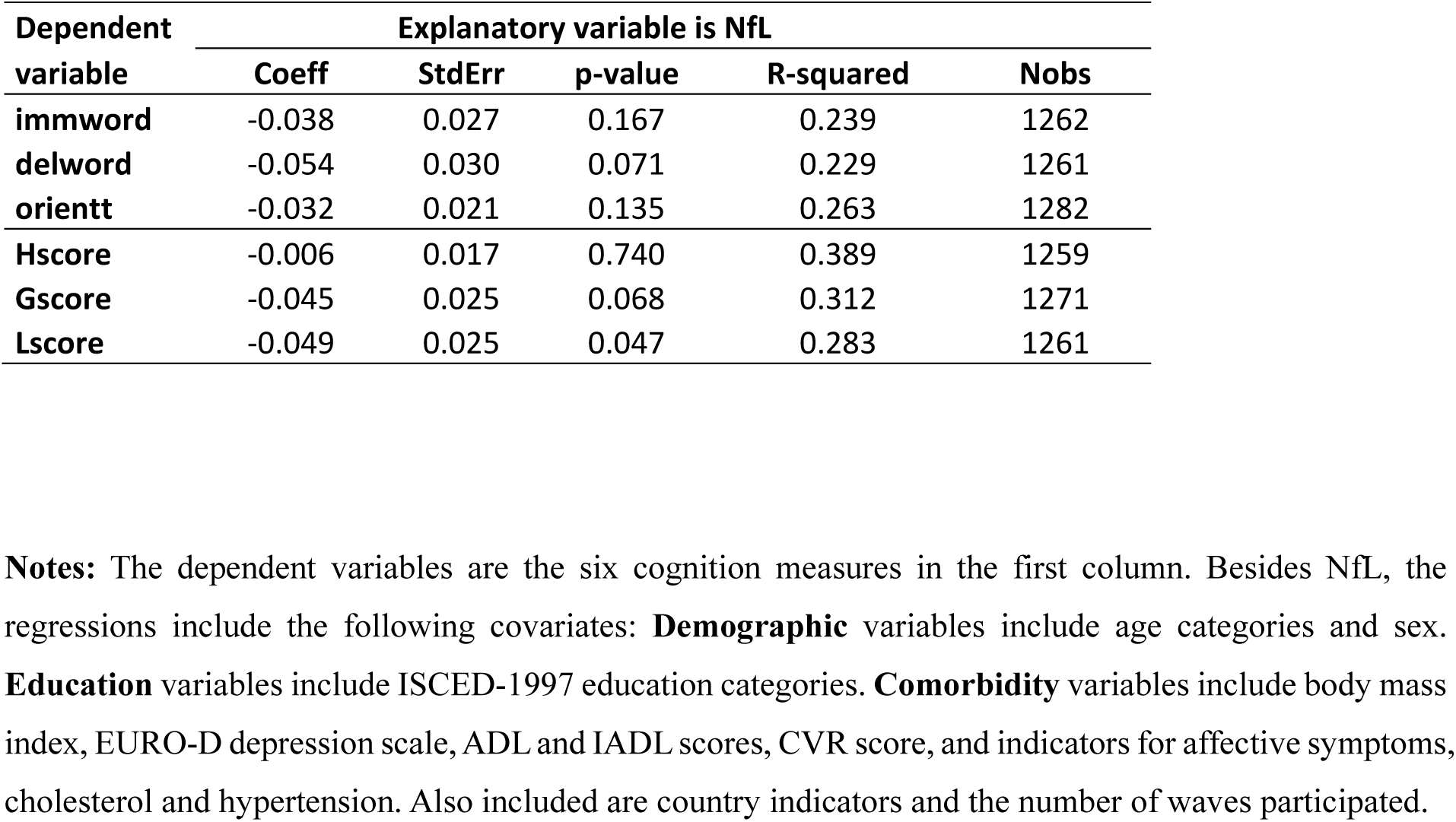
Predictive power of NfL measured seven years earlier.

| Dependent variable | Explanatory variable is NfL |  |  |  |  |
| --- | --- | --- | --- | --- | --- |
|  | Coeff | StdErr | p-value | R-squared | Nobs |
| **immword** | -0.038 | 0.027 | 0.167 | 0.239 | 1262 |
| **delword** | -0.054 | 0.030 | 0.071 | 0.229 | 1261 |
| **orientt** | -0.032 | 0.021 | 0.135 | 0.263 | 1282 |
| **Hscore** | -0.006 | 0.017 | 0.740 | 0.389 | 1259 |
| **Gscore** | -0.045 | 0.025 | 0.068 | 0.312 | 1271 |
| **Lscore** | -0.049 | 0.025 | 0.047 | 0.283 | 1261 |

**Notes:** The dependent variables are the six cognition measures in the first column. Besides NfL, the regressions include the following covariates: **Demographic** variables include age categories and sex. **Education** variables include ISCED-1997 education categories. **Comorbidity** variables include body mass index, EURO-D depression scale, ADL and IADL scores, CVR score, and indicators for affective symptoms, cholesterol and hypertension. Also included are country indicators and the number of waves participated.

Taking all three tables together, we find evidence for associations between the three neurodegenerative biomarkers pTau217, GFAP and NfL and cognitive performance seven years later, most systematically with the three summary scores. However, the sample is too small for a systematic statistical significance.

### 3.4 Joint analyses

We now include all four biomarkers of neurodegeneration jointly in regressions with the three basic cognitive performance measures and the three summary scores. pTau217, GFAP and NfL are standardized to allow for a comparison of the effect sizes. Table 6 shows the results.

**Table 6:**
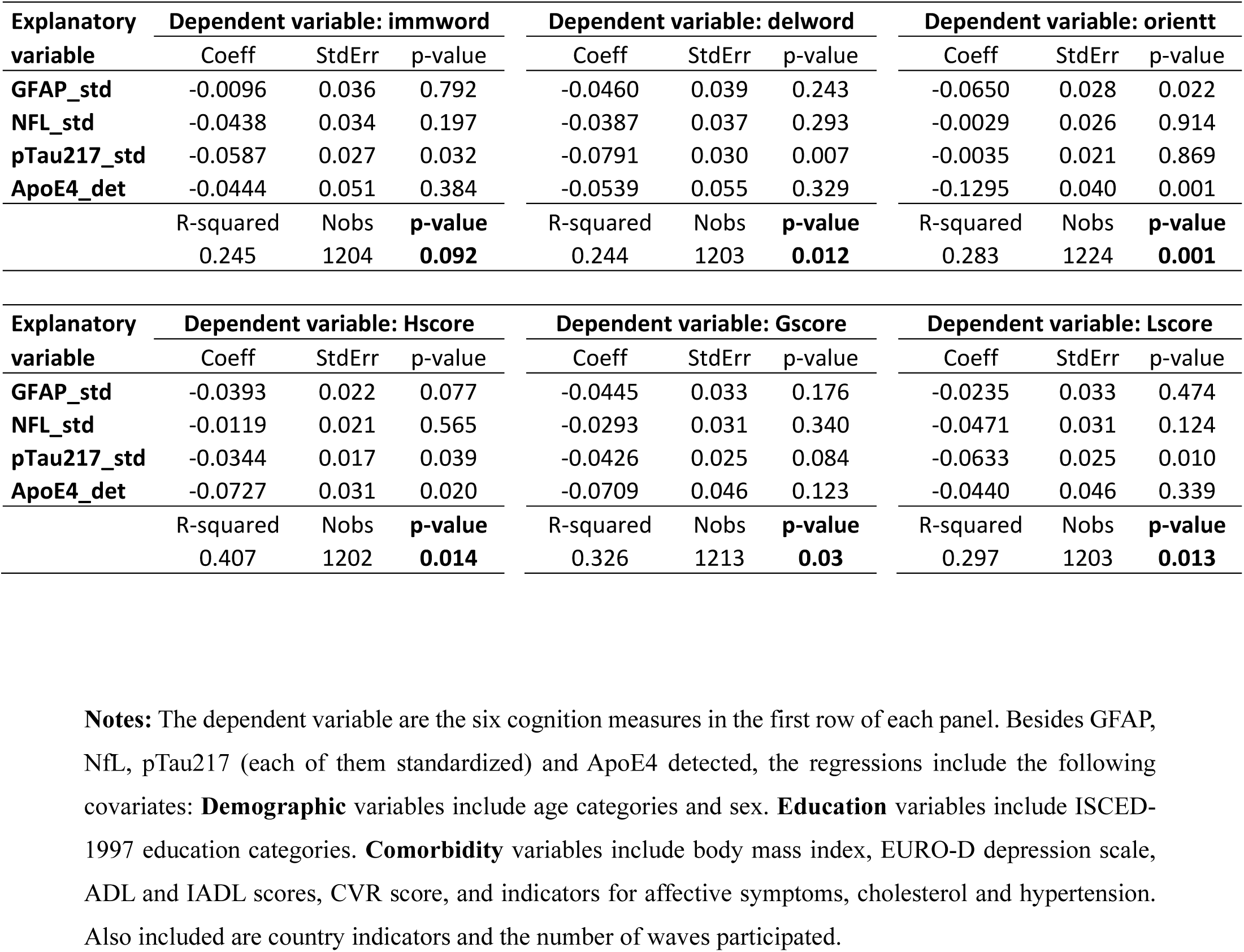
Cognitive performance as function of all four neurodegenerative biomarkers.

| Explanatory variable | Dependent variable: immword |  |  | Dependent variable: delword |  |  | Dependent variable: orientt |  |  |
| --- | --- | --- | --- | --- | --- | --- | --- | --- | --- |
|  | Coeff | StdErr | p-value | Coeff | StdErr | p-value | Coeff | StdErr | p-value |
| GFAP_std | -0.0096 | 0.036 | 0.792 | -0.0460 | 0.039 | 0.243 | -0.0650 | 0.028 | 0.022 |
| NFL_std | -0.0438 | 0.034 | 0.197 | -0.0387 | 0.037 | 0.293 | -0.0029 | 0.026 | 0.914 |
| pTau217_std | -0.0587 | 0.027 | 0.032 | -0.0791 | 0.030 | 0.007 | -0.0035 | 0.021 | 0.869 |
| ApoE4_det | -0.0444 | 0.051 | 0.384 | -0.0539 | 0.055 | 0.329 | -0.1295 | 0.040 | 0.001 |
|  | R-squared | Nobs | p-value | R-squared | Nobs | p-value | R-squared | Nobs | p-value |
|  | 0.245 | 1204 | **0.092** | 0.244 | 1203 | **0.012** | 0.283 | 1224 | **0.001** |

| Explanatory variable | Dependent variable: Hscore |  |  | Dependent variable: Gscore |  |  | Dependent variable: Lscore |  |  |
| --- | --- | --- | --- | --- | --- | --- | --- | --- | --- |
|  | Coeff | StdErr | p-value | Coeff | StdErr | p-value | Coeff | StdErr | p-value |
| GFAP_std | -0.0393 | 0.022 | 0.077 | -0.0445 | 0.033 | 0.176 | -0.0235 | 0.033 | 0.474 |
| NFL_std | -0.0119 | 0.021 | 0.565 | -0.0293 | 0.031 | 0.340 | -0.0471 | 0.031 | 0.124 |
| pTau217_std | -0.0344 | 0.017 | 0.039 | -0.0426 | 0.025 | 0.084 | -0.0633 | 0.025 | 0.010 |
| ApoE4_det | -0.0727 | 0.031 | 0.020 | -0.0709 | 0.046 | 0.123 | -0.0440 | 0.046 | 0.339 |
|  | R-squared | Nobs | p-value | R-squared | Nobs | p-value | R-squared | Nobs | p-value |
|  | 0.407 | 1202 | **0.014** | 0.326 | 1213 | **0.03** | 0.297 | 1203 | **0.013** |

**Notes:** The dependent variable are the six cognition measures in the first row of each panel. Besides GFAP, NfL, pTau217 (each of them standardized) and ApoE4 detected, the regressions include the following covariates: **Demographic** variables include age categories and sex. **Education** variables include ISCED-1997 education categories. **Comorbidity** variables include body mass index, EURO-D depression scale, ADL and IADL scores, CVR score, and indicators for affective symptoms, cholesterol and hypertension. Also included are country indicators and the number of waves participated.

The most important result are the F-tests of joint statistical significance, which are marked bold in the lower right corner of each of the six panels, corresponding to the six cognitive performance measures. All F-tests are significant with the exception of immediate word recall. All signs show a systematic pattern of worsening performance with elevated neurodegenerative biomarker levels. pTau217 has the strongest significance in this joint analysis, and, with few exceptions, also the strongest effect size.

### 3.5 Robustness

As we have demonstrated in Table 2, our findings are robust to a large set of risk factors that might have confounded our results. To assess the robustness of our primary findings further, we conducted several additional analyses. A potential challenge is the large attrition rate in the seven years between Wave 6 and Wave 9, in particular due to the Covid-19 pandemic which disturbed the routine pattern of data collection and created an unusually high non-response rate. Of the 13,102 respondents with valid ApoE4 information in Wave 6, only 6,523 respondents also participated in Wave 9 and had valid cognition information. This corresponds to an attrition rate of 50.2%. Seven years of mortality account for about a third of this attrition, about a similar percentage has been lost due to refusal to either unit non-response or item non-response related to the cognition module, and another third for unknown reasons, mostly mobility with unknown destination. This attrition is most likely not random. One might suspect that respondents with cognition challenges are more likely to drop from the sample. To address this selection issue, we model attrition in a regression parallel to the prediction regression. To simplify the analysis, we use the dichotomized version of Hscore, the HCAP-validated cognition indicator HCI, and performed multinomial logit analyses of the probability to be severely impaired and the probability of attrition. Table 7 shows the results.

**Table 7:**
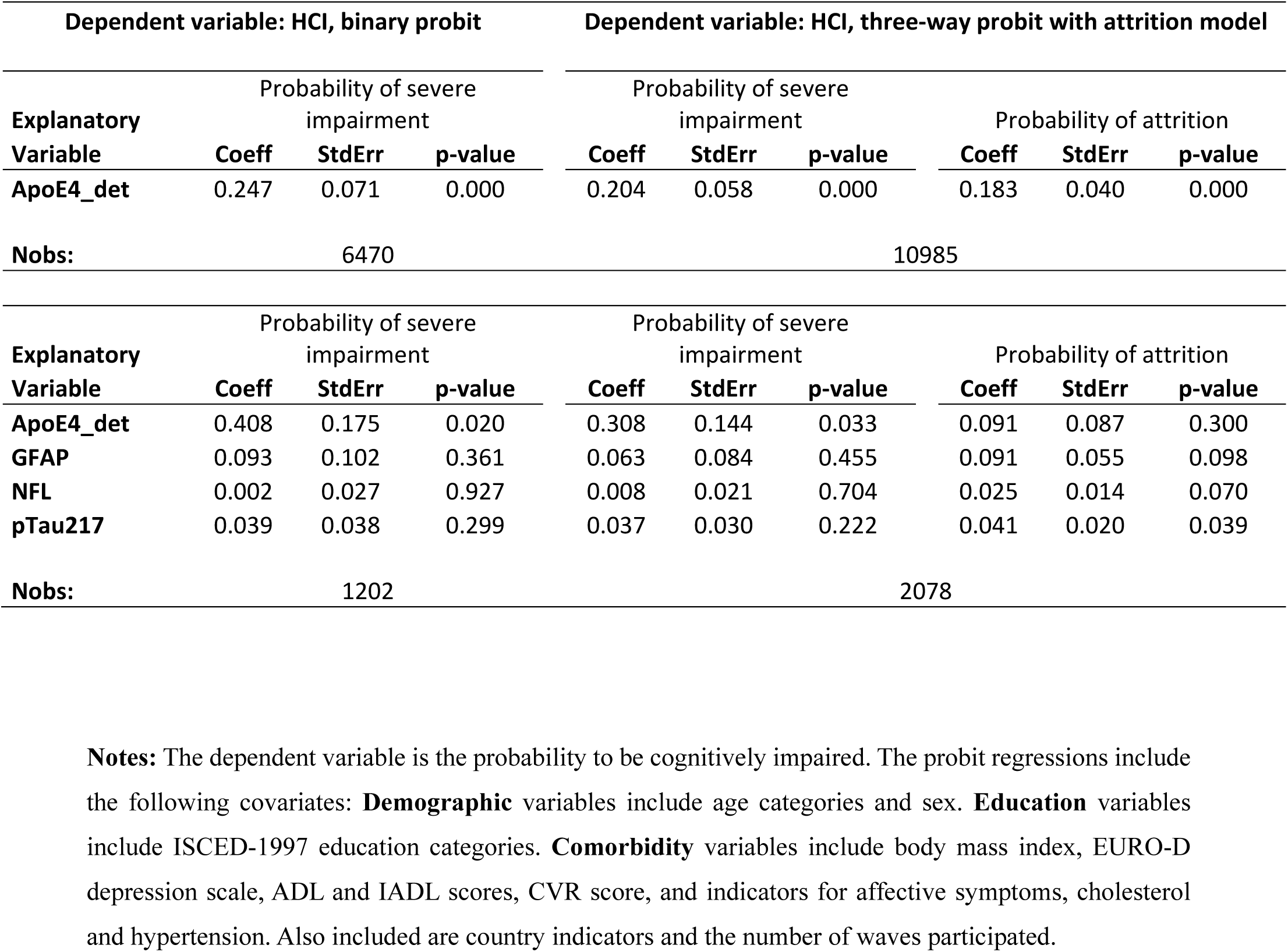
Test for attrition bias.

| Dependent variable: HCI, binary probit |  |  |  | Dependent variable: HCI, three-way probit with attrition model |  |  |  |  |  |
| --- | --- | --- | --- | --- | --- | --- | --- | --- | --- |
| Explanatory Variable | Probability of severe impairment |  |  | Probability of severe impairment |  |  | Probability of attrition |  |  |
|  | Coeff | StdErr | p-value | Coeff | StdErr | p-value | Coeff | StdErr | p-value |
| ApoE4_det | 0.247 | 0.071 | 0.000 | 0.204 | 0.058 | 0.000 | 0.183 | 0.040 | 0.000 |
| Nobs: | 6470 |  |  | 10985 |  |  |  |  |  |
| Explanatory Variable | Probability of severe impairment |  |  | Probability of severe impairment |  |  | Probability of attrition |  |  |
|  | Coeff | StdErr | p-value | Coeff | StdErr | p-value | Coeff | StdErr | p-value |
| ApoE4_det | 0.408 | 0.175 | 0.020 | 0.308 | 0.144 | 0.033 | 0.091 | 0.087 | 0.300 |
| GFAP | 0.093 | 0.102 | 0.361 | 0.063 | 0.084 | 0.455 | 0.091 | 0.055 | 0.098 |
| NFL | 0.002 | 0.027 | 0.927 | 0.008 | 0.021 | 0.704 | 0.025 | 0.014 | 0.070 |
| pTau217 | 0.039 | 0.038 | 0.299 | 0.037 | 0.030 | 0.222 | 0.041 | 0.020 | 0.039 |
| Nobs: | 1202 |  |  | 2078 |  |  |  |  |  |

**Notes:** The dependent variable is the probability to be cognitively impaired. The probit regressions include the following covariates: **Demographic** variables include age categories and sex. **Education** variables include ISCED-1997 education categories. **Comorbidity** variables include body mass index, EURO-D depression scale, ADL and IADL scores, CVR score, and indicators for affective symptoms, cholesterol and hypertension. Also included are country indicators and the number of waves participated.

The left side of Table 7 shows the analysis ignoring attrition while the right side includes a separate equation for attrition, depending on the same explanatory variables. Taking attrition into account attenuates both significance and magnitude of the estimated associations but the essence of our findings remains robust.

A second robustness check examines the consistency of the association between ApoE4 and cognitive impairment across earlier waves of data collection (Waves 8, 7, and 6), in addition to the primary Wave 9 analysis. We focus on Gscore and its dichotomized counterpart (the GCI indicator) since it is available for all four waves (W9, W8, W7, and W6), and the Langa-Weir indicator for Waves 9 and 8, while HCAP-validated cognition information is only available for Wave 9. Results are presented in Supplemental Section S5. For GCI, the association with ApoE4 remains positive in all four waves, suggesting a consistent trend of increased odds of global cognitive impairment among carriers of the gene. The association is statistically significant in Wave 9 and Wave 8. For LWI, a statistically significant positive association with ApoE4 is observed in both Wave 9 and Wave 8. Hence, the general pattern of a strong and negative association between ApoE4 gene and indicators of cognitive impairment hold across multiple time points, further strengthening the robustness of our findings. This robustness holds despite variations in sample size, country composition in Wave 7, and minor differences in covariate adjustment for earlier waves.

## 4. Discussion

This paper documents that four biomarkers of neurodegeneration (ApoE4 protein, pTau217, GFAP, and NfL) collected from dried blood spots in a representative population sample and under normal survey conditions are useful and scalable predictors of cognitive impairment many years later. In the relatively large sample with ApoE4 information, we find that respondents with ApoE4 detection have worse cognitive performance seven years later as measured by all three basic cognitive performance measures and all three cognition summary scores. In the smaller sample with pTau217, GFAP and NfL information, pTau217 has the most systematic association with cognitive performance seven years later, and the combination of all four biomarkers of neurodegeneration is significant for all six cognitive performance measures except immediate word recall. Adding further risk factors into the regression equations takes away variation that is unrelated to the biomarkers, hence it strengthens these associations rather than weakens them due to confounding, suggesting a direct relationship of these proteins with cognitive performance.

Since measuring the level of the biomarkers can be done on the basis of dried blood spots under normal conditions in the community outside of clinical settings, it provides a cost-efficient and scalable early warning signal enabling preventive measures against AD/ADRD before the onset of serious symptoms. This early detection is important since dementia is currently without cure, so initiating preventive measures such as life-style changes early is the key to postpone the onset of dementia and to reduce the burden of AD/ADRD.

The combination of ApoE4 and education explains more than 40 percent of the variation in cognitive performance, even without any other health or behavioral covariate. Largely, the destiny of suffering from dementia later in life is therefore determined early in life. While ApoE4 is genetically given, education is a modifiable risk factor, as are life style and health behaviors. The large predictive power shown in this study gives early detection the power to change this destiny.

In addition to the robustness with respect to the inclusion of a large set of covariates, our results are also robust with respect to the specification of cognitive performance. We use three different summary measures of cognitive impairment, each consisting of several indicators of cognitive performance. We conclude that ApoE4 negatively affects a large spectrum of cognitive tasks. Moreover, we use dichotomized and continuous variants of the three summary measures of cognition, finding similar results. Robustness is also confirmed by augmenting the analysis by an explicit equation that describes attrition between Wave 6 and Wave 9, plus using analyses with different points in time and various sample definitions.

Nevertheless, some caveats remain. The measurement of education by the ISCED code and its attribution to the current country of residence is not perfect as respondents may have moved since childhood. However, respondents with migratory background are less than 3% of the sample. The main challenges of our analysis are the small sample with pTau217, GFAP and NfL information and the long time between collection and analysis of the corresponding DBS samples. While we find sufficient statistical power for a joint analysis of all four biomarkers of neurodegeneration, the sample is too small to pick up separate effects. We cannot quantify how much of the pTau217, GFAP and NfL material has decayed during storage between the collection in 2015 and the laboratory analysis ten years later. In addition, the DBS samples have been stored at −20 °C but freeze/thaw has occurred in 2016 for division of DBS samples into subsamples for the analysis of conventional biomarkers and cytokines, incl. ApoE4. After sample processing, the DBS were returned to the biobank. pTau217, GFAP and NfL from fresh DBS may have been easier to detect even in a relatively small sample. If one wants to shed more light on the long-run predictive properties of pTau217, GFAP and NfL, it may be advisable to collect new DBS samples, preferably in a larger population sample, and analyze them shortly thereafter to establish baseline blood values.

## Data Availability

Access to SHARE data is granted to registered users via the SHARE Research Data Center (http://www.share-project.org/data-access.html).

## Acknowledgments

Funding for the collection and analysis of the DBS data was granted by the US National Institute on Aging (R01 AG063944). Funding for the collection and analysis of the SHARE-HCAP data was granted by the US National Institute on Aging (R01 AG056329). The EU-Commission’s contribution to SHARE-HCAP and the SHARE parent study through the H2020 framework programme (SHAREDEV3, No. 676536) is gratefully acknowledged. Data collection was also supported by national sources in almost all SHARE countries. The funders of the study had no role in study design, data collection, data analysis, data interpretation, or writing of the report.

## Consent Statement

All human subjects in this study provided informed consent.

## Author contributions

ABS conceptualized and designed the SHARE parent study, and did the funding acquisition. ABS and MBS conceptualized and designed the collection of DBS. ABS and SD conceptualized and designed the SHARE-HCAP study. NBL supervised the laboratory analysis of the biomarkers. MCO and SD conducted factor analyses and classification of the HCAP cognition measures. ABS, YDL and YP conducted the DBS data analysis and interpreted the results pertaining to ApoE4. ABS and MCO conducted the DBS data analysis and interpreted the results pertaining to pTau217, GFAP and NfL. ABS and YP wrote the original draft with all co-authors providing review and editing. All authors had full access to the data in the study, verified the data and approved the final version of the manuscript.

## Supplementary material

### Contents

Section S1: Sensitivity of pTau217, GFAP and NFL to survey conditions Section S2: Data preparation

Section S3: Sample characteristics Section S4: Detailed results Section S5: Robustness checks

### Section S1: Sensitivity of pTau217, GFAP and NFL to survey conditions

Table S1 shows means and standard deviations of fieldwork conditions: spot size in centimeters, drying time in minutes, shipment time in days, outside temperature in centigrade, and whether the shipping bag was open or closed:

**Table S1:** Fieldwork conditions.

| Variable | Obs | Mean | Std. Dev. | Min | Max |
| --- | --- | --- | --- | --- | --- |
| t_size | 10,985 | 0.610599 | 0.36074 | .0918805 | 2.082232 |
| t_dry | 10,988 | 21.51119 | 9.32604 | 5 | 40 |
| t_ship | 10,988 | 5.560174 | 3.64595 | 1 | 20 |
| t_temp | 10,988 | 16.36184 | 8.48741 | 0 | 35 |
| t_open | 10,988 | 0.089370 | 0.28529 | 0 | 1 |

We then regressed pTau217, GFAP and NFL in these fieldwork conditions (Table S5). We find a positive association between GFAP and drying time and a negative association between GFAP and outside temperature and whether or not bag was opened before or during shipping. No significant associations were found between Tau217 and NFL on the one hand and the fieldwork conditions on the other hand.

**Table S2:**
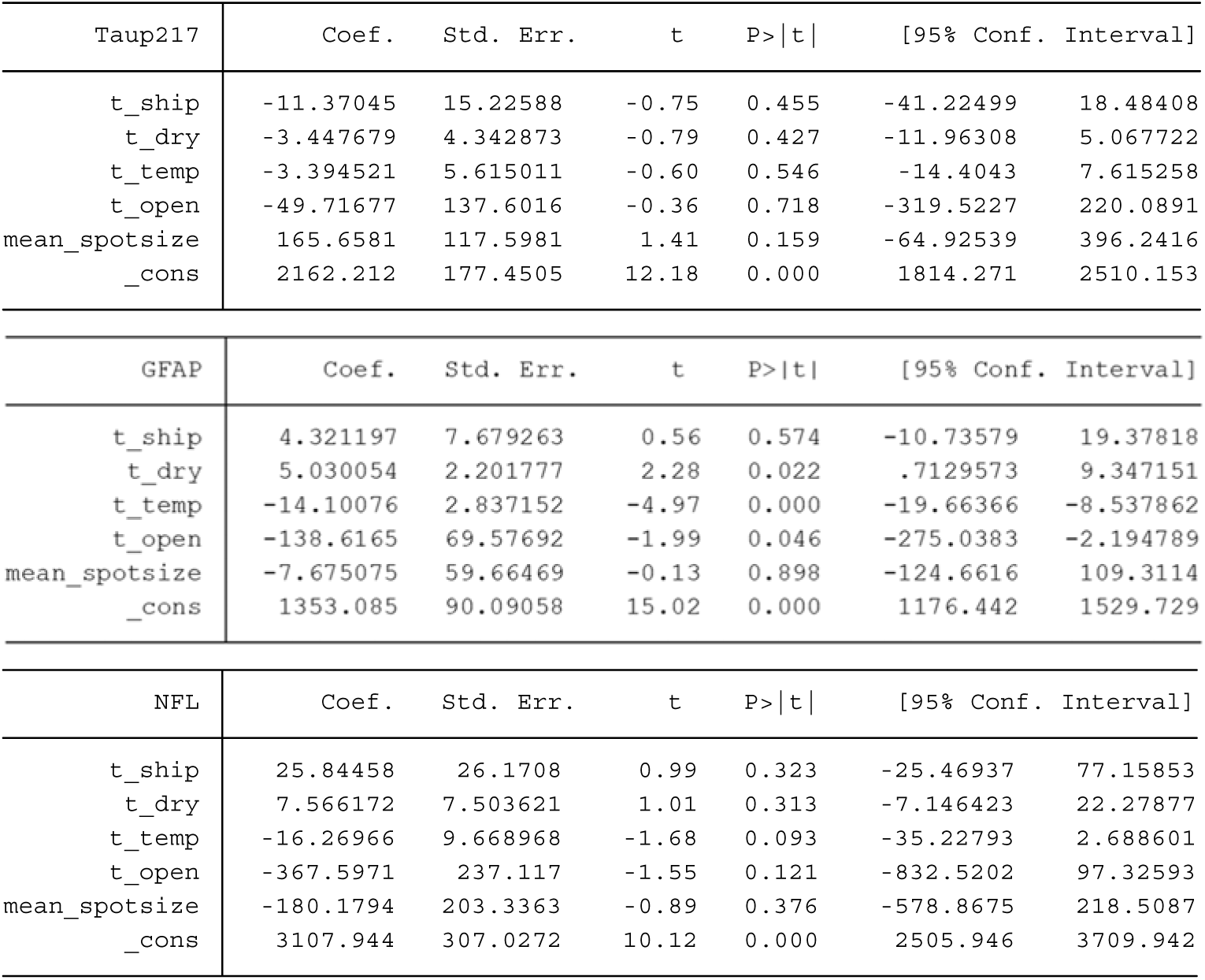
Correlation between pTau217, GFAP and NFL and fieldwork conditions.

| Taup217 | Coef. | Std. Err. | t | P>\|t\| | [95% Conf. Interval] |  |
| --- | --- | --- | --- | --- | --- | --- |
| t_ship | -11.37045 | 15.22588 | -0.75 | 0.455 | -41.22499 | 18.48408 |
| t_dry | -3.447679 | 4.342873 | -0.79 | 0.427 | -11.96308 | 5.067722 |
| t_temp | -3.394521 | 5.615011 | -0.60 | 0.546 | -14.4043 | 7.615258 |
| t_open | -49.71677 | 137.6016 | -0.36 | 0.718 | -319.5227 | 220.0891 |
| mean_spotsize | 165.6581 | 117.5981 | 1.41 | 0.159 | -64.92539 | 396.2416 |
| _cons | 2162.212 | 177.4505 | 12.18 | 0.000 | 1814.271 | 2510.153 |

| GFAP | Coef. | Std. Err. | t | P>\|t\| | [95% Conf. Interval] |  |
| --- | --- | --- | --- | --- | --- | --- |
| t_ship | 4.321197 | 7.679263 | 0.56 | 0.574 | -10.73579 | 19.37818 |
| t_dry | 5.030054 | 2.201777 | 2.28 | 0.022 | .7129573 | 9.347151 |
| t_temp | -14.10076 | 2.837152 | -4.97 | 0.000 | -19.66366 | -8.537862 |
| t_open | -138.6165 | 69.57692 | -1.99 | 0.046 | -275.0383 | -2.194789 |
| mean_spotsize | -7.675075 | 59.66469 | -0.13 | 0.898 | -124.6616 | 109.3114 |
| _cons | 1353.085 | 90.09058 | 15.02 | 0.000 | 1176.442 | 1529.729 |

| NFL | Coef. | Std. Err. | t | P>\|t\| | [95% Conf. Interval] |  |
| --- | --- | --- | --- | --- | --- | --- |
| t_ship | 25.84458 | 26.1708 | 0.99 | 0.323 | -25.46937 | 77.15853 |
| t_dry | 7.566172 | 7.503621 | 1.01 | 0.313 | -7.146423 | 22.27877 |
| t_temp | -16.26966 | 9.668968 | -1.68 | 0.093 | -35.22793 | 2.688601 |
| t_open | -367.5971 | 237.117 | -1.55 | 0.121 | -832.5202 | 97.32593 |
| mean_spotsize | -180.1794 | 203.3363 | -0.89 | 0.376 | -578.8675 | 218.5087 |
| _cons | 3107.944 | 307.0272 | 10.12 | 0.000 | 2505.946 | 3709.942 |

### Section S2: Data preparation

#### S2.1 Data preparation and processing

The data used in the study combines data from the SHARE parent study Waves 6 through 9, the Dried Blood Spots (DBS) data collection in Wave 6, and the Harmonized Cognitive Assessment Protocol (HCAP) data collection in Wave 9. The dataset includes variables derived from the cognitive tests and extrapolated cognitive classifications for SHARE 9 respondents who did not participate in SHARE-HCAP. The main analysis is restricted to individuals who

1. participated in the Dried Blood Spots (DBS) data collection in Wave 6;
2. completed the cognitive module in Wave 9;
3. were born in 1957 or earlier.

Initially, we prepared the raw data for each wave. This step included data cleaning, variable recoding, and the generation of derived variables. The outcome of this phase was a set of structured datasets, ready for subsequent analyses. Next, we proceeded to generate key cognitive impairment indicators. These included the Global Cognitive Impairment (GCI) index, the HCAP Cognitive Impairment index (HCI), and the Langa-Weir Index (LW), all serving as primary outcome variables in our analysis. To ensure a complete dataset, missing values in relevant variables were imputed. This crucial step prepared the data for robust analysis. Finally, we generated attrition weights for each wave, accounting for the complex sampling design inherent in SHARE data. These weights are essential for ensuring the representativeness of our findings.

The following sections describe in detail the operational steps made. This steps refers to wave 9 data. Similar protocol has been applied to wave 8, wave 7 and wave 6 datasets.

#### S2.2 Preparing auxiliary datasets: Social activity data, BMI and imputation data

Our initial Wave 9 data preparation involved creating auxiliary datasets to support later analyses. First, we derived social activity participation from the AC module data. We created a binary indicator, which captured engagement in activities like voluntary work, educational courses, club membership, or political/community involvement over the past year. This variable was based on participation frequency, from daily to less frequent engagement.

Additionally, we used imputed general variable data to extract key variables. These included Body Mass Index (BMI), the Instrumental Activities of Daily Living (IADL) scale, and the EURO-D depression scale. For these, we only kept the first imputation. BMI was used to classify individuals by weight-for-height. The IADL index, adapted for SHARE, quantified limitations in daily living activities, with higher scores indicating greater functional impairment. The EURO-D scale, a 12-item instrument, assessed depressive symptoms, and its scores were dichotomized to indicate clinically relevant depression.

#### S2.3 Selecting and cleaning core variables

We built our core analytical dataset from a comprehensive dataset provided by the DBS and HCAP team at MEA-SHARE. This dataset, which integrated all SHARE waves with Wave 6 DBS data and extrapolated HCAP-validated probabilities for Wave 9 respondents, served as our foundation.

From this base, we selected a subset of variables covering demographic characteristics, cognitive function, health conditions, medication usage, and biomarker data. To focus specifically on Wave 9, we excluded any observations where the Wave 9 sample weights for individuals aged 50 and above were missing.

We then initiated a thorough cleaning process for cognitive function (CF) variables. This involved dropping observations with missing values when interviews were conducted with proxy respondents. We also converted “Don’t know” or “refusal” responses to missing values across cognitive variables, diagnosed diseases, medication usage, and IADL variables. The age variable underwent similar adjustments. Furthermore, we corrected the education variable by recoding “no schooling” (originally 9000) to 0, and we made necessary corrections to the ISCED 1997 education variable.

The cognitive variables used to create the composite score for global cognitive impairment included:

- Verbal episodic memory, assessed by two word recall tests.
- Temporal orientation, determined by the respondent’s knowledge of the current date, month, year, and day of the week.
- Verbal fluency, measured by the number of animals named within 60 seconds.
- Numeracy, evaluated through a serial subtraction task.

Finally, we renamed all variables to ensure consistency and clarity throughout our analyses, removing any wave-specific nomenclature.

#### S2.4 Variable Generation and Recoding

After cleaning the core dataset, we moved on to generating and recoding variables. We started with demographic variables. We created a variable to track the number of waves a participant completed and a binary indicator for gender. We also generated country labels and established a set of age category variables: 5-year age spans starting from age 65.

Next, we derived physical health variables. We created a binary variable to represent physical activity engagement, categorizing responses as either “once a week or more” or “less than once a week.” We also calculated variables for Mini-Mental State Examination (MMSE) components (temporal orientation), Instrumental Activities of Daily Living (IADL) scores, and Cardiovascular Risk (CVR) scores. The CVR index was specifically computed based on the presence of stroke, myocardial infarction, and diabetes.

For biomarker data, we created a dummy variable (DBS) to indicate the presence of the ApoE4 genotype.

Finally, we processed information related to self-declared medical diagnoses and medication intake. This involved capturing whether respondents reported a medical diagnosis and/or were currently undergoing treatment for a specific list of conditions, and whether they took any medication at least once a week.

#### S2.5 Selecting relevant variables

To focus our analysis, we began by selecting a specific set of variables directly relevant to our research questions. These included: demographic variables (age, gender, country), education, number of waves, biomarker data (ApoE4), cognitive measures (recall, MMSE, fluency, subtraction), health conditions (BMI, EuroD, chronic diseases), medication usage, activity variables (physical activity, social activity), drinking and smoking behavior, Langa-Weir variables, the cognitive class variable, and weight variables.

Subsequently, we refined our sample to include only individuals with complete information for the ApoE4 protein. We retained participants exclusively if their ApoE4 genotype was recorded, as this measure is central to our study.

#### S2.6 Generating the cognitive impairment indicators

To measure cognitive impairment, we used several different methods. For the first method, called Global cognition score (Gscore), we looked at five specific cognitive tests (MMSE, two recall tests, a subtraction test, and a fluency test). We counted how many of these tests were missing for each person, and also how many were missing overall. Then, we used a statistical technique to compare each person’s test scores to the average scores. This gave us a ’Z-score’ for each person on each test. We then averaged these Z-scores to get a single ’Z-mean’ for each person, but we only did this if they had at least two test scores. If someone was missing too many tests, we didn’t calculate a Z-mean for them. Following a method used in a previous study (Deza-Lougovski et al.), we then decided who had cognitive impairment based on their Z-mean. We defined as cutoff point one standard deviation below mean, and anyone whose Z-mean was below this cutoff was considered to have cognitive impairment. This indicator is named GCI.

Secondly, we employed the Langa-Weir approach, as detailed in Crimmins (2011), to classify cognitive impairment. This method, originally developed using data from the Health and Retirement Study (HRS), aims to identify individuals with dementia and those with Cognitively Impaired Not Demented (CIND). Due to limitations in data availability for participants under 65 years old, the Langa-Weir method in this study utilized a subset of cognitive measures. These included immediate and delayed recall, serial 7s subtraction, and backward counting, resulting in a composite score ranging from 0 to 27. Based on this score (Lscore), self-respondents were categorized as follows: ’Demented’ (scores 0-6), ’CIND’ (scores 7-11), and ’Normal’ (scores 12-27). For our analysis, we created a binary variable LWI to denote cognitive impairment, where 1 indicates either ’Demented’ or ’CIND’ and 0 indicates ’Normal’.

Finally, we used the probabilities of being in different cognitive states to construct the HCAP-validated cognition score (Hscore). These probabilities were computed from regression analyses based on the HCAP subsample on the basis of 27 detailed cognition tests and then extrapolated to the entire SHARE W9 sample using all common cognition-related variables that were contained in both the HCAP subsample and the SHARE Wave 9 parent sample (Börsch-Supan et al. 2025). We also created a new category variable HCI to classify people based on these probabilities. If a person’s probability of being in the first cognitive state was higher than their probability of being in the second or third states, we assigned them to category 1. We did the same thing for categories 2 and 3.

Essentially, we assigned people to the cognitive state they were most likely to be in, according to these probabilities. Gscore, Lscore and Hscore were then standardized to mean zero and standard deviation one.

#### S1.7 Attrition weights

Individuals who continue to participate in longitudinal studies may differ systematically from those who discontinue their involvement. By computing attrition weights, we aimed to correct for such non-random attrition, thereby strengthening the validity and generalizability of our analyses.

The dataset used for constructing the weights was derived from a comprehensive dataset encompassing all waves of the SHARE study. From this source, we extracted a selection of variables, including unique identifiers, participation status in relevant waves, demographic characteristics, educational attainment, country of residence, and initial Wave 6 DBS weights. This subset was then integrated with our Wave 9 analytical sample, specifically restricted to individuals aged 65 and above who had a non-missing cognitive score. The merge was configured to retain only those observations present in both datasets.

Propensity scores for inclusion in the Wave 9 sample, conditional on Wave 6 participation, were estimated using logistic regression. This was done separately for each cognitive impairment (CI) indicator: HCI, GCI, and LWI. These propensity scores were derived from unweighted Wave 6 observations. For any observations not present in the Wave 9 sample, their propensity scores were set to missing. New weights were then calculated as the inverse of these estimated propensity scores. This process generated a specific set of weights for each CI indicator, directly corresponding to their respective unweighted propensity scores.

#### S1.8 Missing values

Table S1 presents the extent of missing data across the variables of interest. Starting with the sample of 6,678 Wave 9 (W9) participants who also participated in the Wave 6 (W6) DBS collection and for whom a valid ApoE4 protein measure was available, the number of missing values for each variable is detailed. A threshold of 25 missing cases was established to guide the decision to employ SHARE imputed values, specifically for variables where these imputed data were available. Consequently, only missing values for BMI, ADL, and IADL scores were imputed using SHARE provided imputation datasets. For EuroD, a new dummy was created to indicate the missing values. For all other variables, observations with missing values were excluded from the final analytical sample

**Table S3:** Missing values

| Variable | Missing | Action |
| --- | --- | --- |
| HCI | 206 | EfS |
| GCI | 155 | EfS |
| LWI | 202 | EfS |
| Detected apoE4 | 0 | None |
| Female | 0 | None |
| Age | 0 | None |
| Education | 0 | None |
| Number waves participated | 0 | None |
| Country | 0 | None |
| BMI | 144 | Imputed |
| Euro-D | 103 | Extra category |
| ADL score | 26 | Imputed |
| IADL score | 26 | Imputed |
| Affective | 18 | EfS |
| CVR risk score | 18 | EfS |
| High cholesterol | 18 | EfS |
| Hypertension | 18 | EfS |

**Notes:** The first column presents the variables employed in the analysis. The second column reports the frequency of missing values for each respective variable. The third column details the methodology adopted for handling these missing values: "EfS - Excluded from Sample" indicates that observations with missing values for the specified variable were excluded from the analysis; "Imputed" signifies that missing values were recovered from the first imputation of the dataset provided by SHARE; "Extra category" denotes the creation of a dummy variable to represent observations with missing values for that variable; "None" indicates that no specific action was required for that variable.

To assess the implications of excluding observations with missing values, Table S2 presents the means and standard deviations for the cognitive impairment indicators across three distinct samples:

A. observations with only complete, non-missing data;
B. observations including imputed values; and
C. observations excluded from the analysis due to missing data.

The analytical sample comprises samples (A) and (B). Despite the relatively small sample size of the excluded observations (C), the higher mean values observed for all three cognitive impairment indicators (HCI, GCI, and LWI) suggest a greater concentration of individuals with cognitive impairment among those with missing data. The inclusion of sample (B) in the analytical sample serves to mitigate the potential loss of information associated with the exclusion of cases with missing values.

**Table S4:** Distribution of cognitive impairment indicators over different samples.

|  |  | Sample |  |  |  |  |  |
| --- | --- | --- | --- | --- | --- | --- | --- |
|  |  | A | B | C | A+B+C | D | A+B+C+D |
| **HCI** | N | 6191 | 228 | 53 | 6472 | 206 | 6678 |
|  | Mean | 0.10 | 0.43 | 0.71 | 0.12 |  |  |
|  | sd | 0.31 | 0.49 | 0.45 | 0.33 |  |  |
| **GCI** | N | 6191 | 228 | 104 | 6523 | 155 | 6678 |
|  | Mean | 0.05 | 0.35 | 0.49 | 0.07 |  |  |
|  | sd | 0.22 | 0.48 | 0.50 | 0.25 |  |  |
| **LWI** | N | 6191 | 228 | 57 | 6476 | 202 | 6678 |
|  | Mean | 0.16 | 0.53 | 0.77 | 0.18 |  |  |
|  | sd | 0.37 | 0.49 | 0.42 | 0.39 |  |  |

**Notes:** HCI is HCAP-validated cognitive impairment indicator, GCI Global cognitive impairment indicator, and LWI Langa-Weir indicator of cognitive impairment. Sample A comprises observations with complete, non-missing data. Sample B consists of observations including imputed values. Sample C includes observations excluded from the analysis due to missing data. Sample D represents observations with missing values for the aforementioned cognitive impairment indicators. "A+B+C" reports the mean and standard deviation for the aggregate sample with valid measurements for the cognitive impairment indicators.

### Section S3: Sample characteristics

Table S5 presents the sample means and standard deviations for all variables used in the paper, based on our analytical sample with 6,523 observations.

**Table S5:** Sample characteristics.

| Variables | Mean | SD | Variables | Mean | SD |
| --- | --- | --- | --- | --- | --- |
| Cognitive indicators |  |  | Health |  |  |
| HCI [0/1] | 0.12 |  | BMI | 26.8 | (4.8) |
| GCI [0/1] | 0.06 |  | EuroD = No | 0.70 |  |
| LWI [0/1] | 0.18 |  | EuroD = Yes | 0.28 |  |
| Hscore | 0.73 | (0.2) | EuroD = Missing | 0.02 |  |
| Gscore | 0.01 | (0.7) | ADL | 0.23 | (0.78) |
| Lscore | 15.33 | (4.5) | IADL | 0.55 | (1.43) |
| Immediate word recall | 5.1 | (1.8) | Affective | 0.06 |  |
| Delayed word recall | 3.7 | (2.1) | CVR score | 0.33 | (0.57) |
| Orientation in time | 0.16 |  | Cholesterol | 0.31 |  |
| Biomarker |  |  | Hypertension | 0.56 |  |
| Detected ApoE4 | 0.26 |  | Country |  |  |
| pTau217 | 2.08 | (1.8) | Germany | 0.10 |  |
| GFAP | 1.35 | (1.0) | Belgium | 0.14 |  |
| NFL | 3.33 | (4.3) | Estonia | 0.20 |  |
| Demographics |  |  | Spain | 0.03 |  |
| Female | 0.59 |  | Greece | 0.03 |  |
| Age 58-64 | 0.31 |  | Italy | 0.06 |  |
| Age 65-69 | 0.20 |  | Switzerland | 0.09 |  |
| Age 70-74 | 0.15 |  | France | 0.01 |  |
| Age 75-79 | 0.14 |  | Slovenia | 0.08 |  |
| Age 80-84 | 0.12 |  | Denmark | 0.14 |  |
| Age 85+ | 0.09 |  | Israel | 0.01 |  |
| Education |  |  | Sweden | 0.11 |  |
| No School | 0.02 |  | N of waves | 4.92 | (1.20) |
| Primary School | 0.11 |  |  |  |  |
| Lower Secondary School | 0.16 |  |  |  |  |
| Higher Secondary School | 0.33 |  |  |  |  |
| Some College | 0.08 |  |  |  |  |
| College | 0.29 |  |  |  |  |
| Post-College | 0.01 |  |  |  |  |

### Section S4: Detailed results

**Table S6:** Complete regression results for. Table 1**, using Hscore as example**

| Dependent variable is Hscore |  |  |  |  |  |  |  |
| --- | --- | --- | --- | --- | --- | --- | --- |
| Variable | Coeff | StdErr | p-value | Variable | Coeff | StdErr | p-value |
| ApoE4 detected | -0.0678 | 0.0184 | 0.000 | Belgium | -0.0058 | 0.0356 | 0.871 |
| 58-59 | 0.3187 | 0.0415 | 0.000 | Estonia | -0.1473 | 0.0341 | 0.000 |
| 60-64 | 0.2698 | 0.0350 | 0.000 | Spain | -0.5158 | 0.0597 | 0.000 |
| 65-69 | 0.2023 | 0.0346 | 0.000 | Greece | -0.3914 | 0.0614 | 0.000 |
| 70-74 | 0.2155 | 0.0347 | 0.000 | Italy | -0.1538 | 0.0460 | 0.001 |
| 75-80 | 0.1113 | 0.0363 | 0.002 | Switzerland | -0.0487 | 0.0400 | 0.223 |
| 80+ |  |  |  | France | -0.0378 | 0.0765 | 0.622 |
| Female | 0.1415 | 0.0173 | 0.000 | Slovenia | -0.1867 | 0.0402 | 0.000 |
| No education |  |  |  | Denmark | -0.0612 | 0.0354 | 0.084 |
| Primary education | 0.5200 | 0.0720 | 0.000 | Israel | -0.5303 | 0.1023 | 0.000 |
| Lower Sec | 0.5319 | 0.0720 | 0.000 | Sweden | -0.0053 | 0.0372 | 0.886 |
| Higher Sec | 0.6785 | 0.0712 | 0.000 | Spot size | 0.0033 | 0.0210 | 0.876 |
| Some College | 0.7304 | 0.0769 | 0.000 | Drying time | 0.0011 | 0.0009 | 0.220 |
| College | 0.6755 | 0.0716 | 0.000 | Shipping time | -0.0056 | 0.0032 | 0.077 |
| Post-College | 0.7296 | 0.1182 | 0.000 | Ambient temperature | 0.0000 | 0.0012 | 0.977 |
| BMI | 0.0067 | 0.0018 | 0.000 | Bag open | -0.0160 | 0.0284 | 0.574 |
| Euro-D | -0.0074 | 0.0041 | 0.074 | 1-2 waves | -0.0287 | 0.0540 | 0.596 |
| Number ADLs | 0.0303 | 0.0134 | 0.023 | 4+ waves | 0.0168 | 0.0204 | 0.411 |
| Number IADLs | -0.4578 | 0.0079 | 0.000 | Constant | -0.7121 | 0.0975 | 0.000 |
| Affective disorder | -0.0136 | 0.0353 | 0.700 |  |  |  |  |
| Cardiovascular risk | -0.0271 | 0.0171 | 0.112 |  |  |  |  |
| Cholesterol | 0.0037 | 0.0195 | 0.849 | R-squared: | 0.579 |  |  |
| Hypertension | 0.0127 | 0.0178 | 0.477 | Nobs: | 6470 |  |  |

### Section S5: Robustness checks

**Table S7:** Effect between detection of apoE4 specific binding and cognitive status over time. *Regression results (odd ratios) of binary logit models.*

|  | (1)<br>GCI W9 | (2)<br>GCI W8 | (3)<br>GCI W7 | (4)<br>GCI W6 | (5)<br>LWI W9 | (6)<br>LWI W8 |
| --- | --- | --- | --- | --- | --- | --- |
| Gene Detected 0/1 | 1.47*<br>1.1,2.0 | 1.55**<br>1.1,2.1 | 1.26<br>0.8,2.0 | 1.29<br>1.0,1.7 | 1.49***<br>1.2,1.8 | 1.47***<br>1.2,1.8 |
| N | 6503 | 5582 | 2502 | 7782 | 6460 | 5549 |
| Pseudo R2 | 0.37 | 0.40 | 0.43 | 0.34 | 0.32 | 0.32 |
| Log Likelihood | -4618.67 | -4221.62 | -2053.32 | -3699.84 | -8628.03 | -8288.41 |
| Education | x | x | x | x | x | x |
| Demographic | x | x | x | x | x | x |
| Health | x | x | x | x | x | x |

**Notes:** The dependent variables in these models are: (1-4) the Global Cognitive Impairment (GCI) indicator in wave 9 (W9), wave 8 (W8), wave 7 (W7), wave 6 (W6); and (5-6) the Langa-Weir (LWI) indicator in wave 9 (W9) and in wave 8 (W8). Each model is estimated using a sample of individuals aged 65 and over who had non-missing values for the respective cognitive impairment indicator at the respective wave. To account for potential attrition between Wave 6 and the respective wave, all observations are weighted by the inverse of their probability of being present in both waves. The reported coefficients are odds ratios, with 95% confidence intervals presented in brackets. \* $p < 0.05$ ; \*\* $p < 0.01$ ; \*\*\* $p < 0.001$ .
**Education** variables include ISCED-1997 education level (dummies). **Demographic** variables include gender (dummy), age group (dummies), country (dummies), and number of waves. **Health** variables include body mass index (BMI), EURO-D depression scale (dummies), ADL and IADL scores, affective symptoms (dummy), CVR score, cholesterol (dummy), and hypertension (dummy).
It is important to note some specific characteristics of the data collection in earlier waves, particularly for Wave 7. The sample size for the Wave 7 GCI analysis is smaller compared to other waves due to the study design, where a significant portion of panel respondents participated in SHARELIFE, a retrospective life history interview, rather than the standard Wave 7 questionnaire. Furthermore, the set of countries included in the Wave 7 data differs somewhat from those in other waves, which could potentially introduce some heterogeneity. Additionally, due to variations in questionnaire design across waves, two health-related regressors could not be consistently included in the models for Waves 6 and 7, potentially leading to slightly different covariate adjustments in those specific analyses.

